# Ethnic inequities in the patterns of Personalised Care Adjustments for ‘Informed Dissent’ and ‘Patient Unsuitable’: A retrospective study using Clinical Practice Research Datalink

**DOI:** 10.1101/2022.09.30.22280554

**Authors:** B. Hayanga, M. Stafford, M. Ashworth, J. Hughes, L. Bécares

## Abstract

**Objectives:** To examine patterns of PCA reporting for ‘informed dissent’ and ‘patient unsuitable’, how they vary by ethnic group, and whether ethnic inequities can be explained by socio-demographic factors or comorbidities.

**Design:** A retrospective study using routinely collected electronic health records.

**Setting:** Individual patient data from Clinical Practice Research Datalink collected from UK general practice.

**Participants:** Patients with at least one of the 12 Quality and Outcomes Framework (QOF) conditions which had PCA coding options from a random sample of 690,00 patients aged 18+ years on the 1^st^ of Jan 2016.

**Main outcomes measures:** The associations between ethnicity and two PCA reasons (‘Informed Dissent’ and ‘Patient Unsuitable’) were examined using logistic regressions after adjustment for age, sex, multiple QOF conditions and area-level deprivation.

**Results:** The association between ethnicity and the two PCA reasons were in opposite directions. After accounting for age, gender, multiple QOF conditions and area-level deprivation, people of Bangladeshi [OR: 0.69, 95% CI: 0.55 to 0.87], Black African [OR: 0.70, 95% CI: 0.61 to 0.81], Black Caribbean, OR: 0.67, 95% CI: 0.58 to 0.76], Indian [OR: 0.74, 95% CI: 0.66 to 0.83], mixed [OR: 0.86, 95% CI: 0.74 to 0.99], other Asian [OR: 0.74 95% CI: 0.64 to 0.86] and other ethnicity [OR: 0.66, 95% CI: 0.55 to 0.80] were less likely to have a PCA record for ‘informed dissent’ than people of white ethnicity. Only people of Indian ethnicity were significantly less likely than people of white ethnicity to have a PCA record for ‘patient unsuitable’ in fully adjusted models [OR: 0.80, 95% CI: 0.67 to 0.94]. We found ethnic inequities in PCA reporting for ‘patient unsuitable’ among people of Black Caribbean, Black other, Pakistani, and other ethnicity, but these attenuated after adjusting for multiple QOF conditions and/or area level deprivation.

**Conclusion:** Study findings counter the narratives that suggest that people from minoritised ethnic groups often refuse medical intervention. They illuminate the complex relationship between ‘informed dissent’ and (dis)empowerment which requires further scrutiny. They also show ethnic inequalities in PCA reporting for ‘patient unsuitable’ that are linked to clinical and social complexity and should be tackled to improve health outcomes for all.

## BACKGROUND

The Quality and Outcomes Framework (QOF), is a pay-for-performance scheme in England which rewards practices for the delivery of evidence-based standards of care. ^1, 2^ To safeguard patients from inappropriate care and/or prevent practices from being penalised for not achieving targets for reasons beyond their control, the scheme allows general practitioners (GPs) to make personal care adjustments (PCAs), to exclude patients from performance indicators. ^2, 3^ Patients can be exempt from performance targets if a service is unavailable or if a patient is newly registered, newly diagnosed, unsuitable for treatment, does not respond to offers of care or refuses treatment (informed dissent).^3^ Exemption from quality indicators is associated with health outcomes including higher mortality risk and poor control of risk factors. ^4-6^

PCA rates vary between practices, across quality indicators, health conditions and reasons for exemption. Exemptions are more prevalent in disadvantaged groups.^4^ Higher rates of informed dissent are found in practices that have a high number of registered patients, in socioeconomically deprived catchments, and failing to secure maximum renumeration in the previous year. ^7^ Patients who are older, have multiple long-term conditions and live in deprived areas are more likely to be removed from achievement calculations for being unsuitable or because of informed dissent.^6^ People from minoritised ethnic groups are overrepresented in domains that are more likely to be exempted from QOF indicators. Minoritised ethnic groups (with the exception of people of Indian, Chinese, white Irish and white other ethnicity) are more likely to be living in the most overall deprived 10% of neighbourhoods in England in 2019.^8^ Older people from minoritised ethnic groups (with the exception of Black African men and Chinese people) have as many or more long-term conditions as people of white British ethnicity.^9^ However, the evidence from the few studies that have explored ethnic variations is unclear. One study, focused on patients with diabetes, reports that Black and South Asian patients have higher odds of being excluded from the HbA1c indicator when compared to their white counterparts. ^4^ An ecological analysis found lower rates of PCA among minoritised ethnic group people with asthma when compared to their white counterparts.^1^

The aim of this study is to assess whether there are ethnic inequities in PCA reporting. We focus on two PCA reasons: ‘patient unsuitable’ (exempted by a GP for a range of reasons including failure to respond to maximum dose of treatment, adverse reaction to treatment, extreme frailty or supervening condition^17,18^) and ‘informed dissent’ (where patients do not agree to treatment or medical investigation). This will give us insight into the clinical judgments made by GPs and patient choice. Identifying groups that are not included in the QOF scheme can inform strategies to ensure all who are eligible receive recommended standards of care.

## METHODS

### Data sources and population

We conducted a retrospective cohort study using data from the Clinical Practice Research Datalink (CPRD) Aurum which contains longitudinal routinely collected electronic health records from patients in general practice.^10^ CPRD Aurum is representative of the population in England in terms of geographical spread, area-level deprivation, age and gender. In March 2022, CPRD Aurum had approximately 13 million patients who were registered at currently contributing practices.^11^ We analysed CPRD Aurum data linked to ONS data to allow for measurement of area-level deprivation using the Index of Multiple Deprivation (IMD) and Hospital Episode Statistics (HES) to improve completeness of ethnicity recording.^12^ We drew upon a random sample of 690,00 patients aged 18 years and above on the 1^st^ of Jan 2016.

### Measures

We extracted ethnic identity from SNOMED codes recorded by the GP. When ethnicity data was missing/incomplete, we drew this from the HES records. We used the England 2011 census to define ethnic group categories but we combined white British, white Irish and other white because these separate categories were not available in HES. Men, people aged under 45 years, and those with one QOF condition were over-represented in those with missing ethnicity data (Supplementary Table 1). We included all QOF long-term conditions for which there were the option of the two PCA codes of interest; ‘informed dissent’ and ‘patient unsuitability’ (Supplementary Table 2). We identified relevant PCA codes between 1^st^ January 2016 and 31^st^ December 2019 for all patients with the aforementioned QOF conditions. Socioeconomic deprivation was derived from the IMD quintiles based on the lower super output area boundaries of the patient’s address (quintile 1 represents the least deprived). ^12^

### Statistical analysis

We created an analytical sample which included only those with at least one of the 12 QOF conditions at baseline (1^st^ January 2016) and complete data on age, gender, ethnicity, and area-level deprivation. We built separate logistic regression models for each outcome and included i) each covariate separately (Model 1), ii) ethnicity, age and gender (Model 2), iii) model 2 plus multiple QOF conditions (Model 3), iv) model 3 plus IMD (Model 4). We also conducted a sensitivity analysis by creating separate models for men and women. We used RStudio (R04.2.0) for all our analyses. ^13^

## RESULTS

250461 patients had at least one QOF condition and complete data on ethnicity, gender, IMD score and age (Figure 1). The majority were of white ethnicity (89.2%) (Table 1 and Supplementary table 3). The Black Caribbean ethnic group had the highest proportion of people aged 75 and above (40.9%). Approximately 50% of people from Bangladeshi, mixed and Black other ethnic group were under 45 years of age (Supplementary Table 3). Over 40% of people from Bangladeshi, Black African, Black Caribbean, Black other, and Pakistani ethnic groups were living in the most deprived IMD quintile (Table 1).

**Figure 1.**
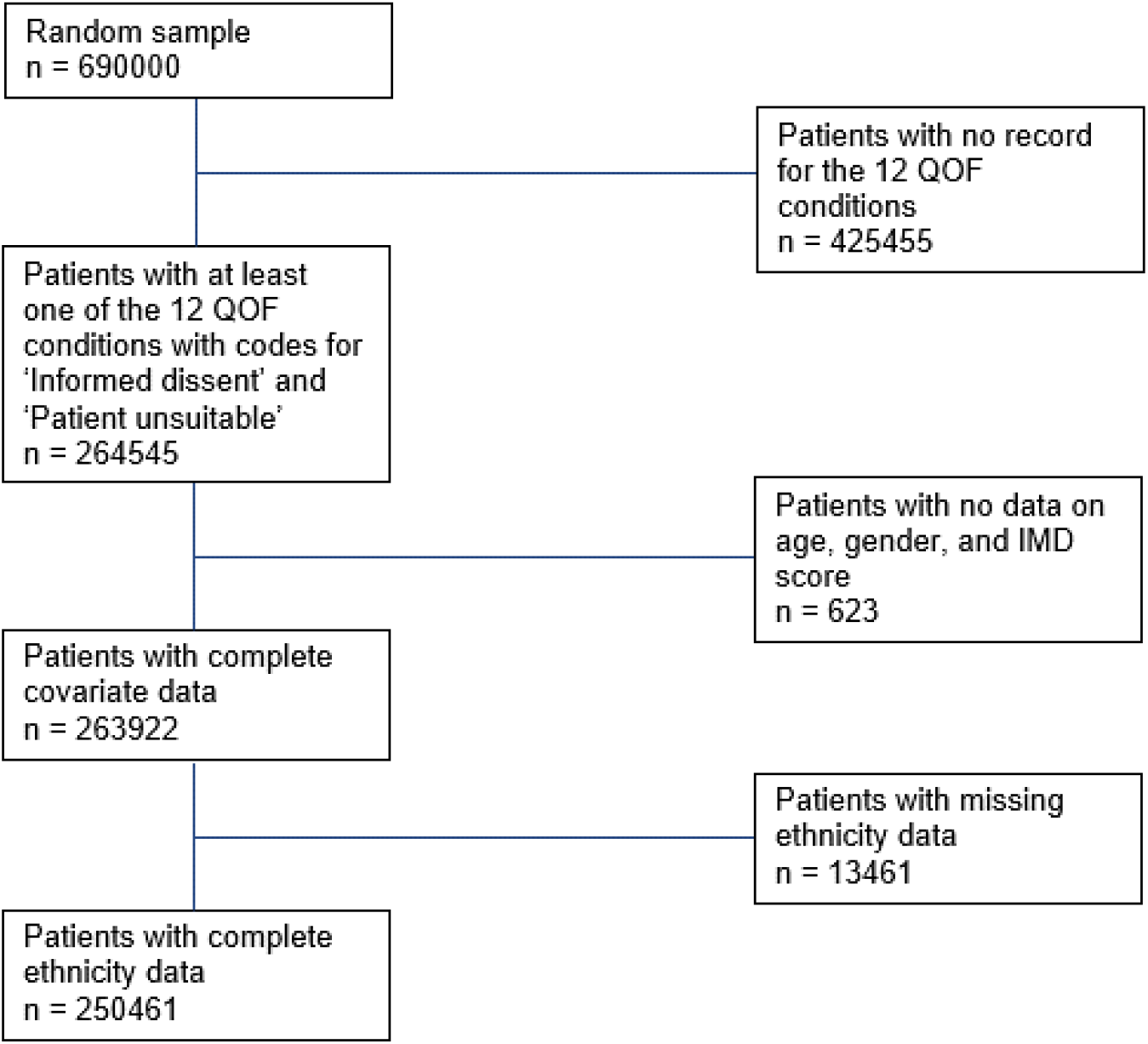
Flow chart to get analytical sample

**Table 1.** Sociodemographic characteristics and percentage of PCA by ethnic group.

|  | n | % Women | % over 75 years | % with multiple QOF conditions | % living in IMD 5 | % with Informed Dissent recorded | % with Patient Unsuitable recorded |
| --- | --- | --- | --- | --- | --- | --- | --- |
| Bangladeshi | 1253 | 49.8 | 7.6 | 32.5 | 50.4 | 6.5 | 3.4 |
| Black African | 3652 | 53.3 | 6.2 | 26.0 | 45.8 | 6.2 | 3.0 |
| Black Caribbean | 3737 | 56.5 | 22.0 | 40.9 | 42.7 | 6.1 | 4.4 |
| Black Other | 1162 | 54.0 | 6.1 | 26.1 | 41.5 | 7.3 | 3.5 |
| Chinese | 741 | 56.4 | 12.6 | 24.2 | 16.5 | 6.5 | 2.0 |
| Indian | 5309 | 50.9 | 13.3 | 38.2 | 19.2 | 6.5 | 2.9 |
| Mixed | 2900 | 56.3 | 7.0 | 25.0 | 30.7 | 7.0 | 2.8 |
| Other Asian | 3035 | 51.0 | 8.7 | 31.2 | 18.8 | 6.3 | 3.1 |
| Other | 2154 | 49.8 | 10.0 | 25.2 | 26.5 | 5.4 | 3.5 |
| Pakistani | 3221 | 49.9 | 8.2 | 34.7 | 40.9 | 8.9 | 3.4 |
| White | 223297 | 54.6 | 20.6 | 36.0 | 17.5 | 8.0 | 3.7 |

Across all ethnic groups, patients were more likely to receive a PCA record for ‘informed dissent’ than for ‘patient unsuitable’. Whilst Pakistani people had the highest proportions of people with a PCA record for ‘informed dissent’ (8.9%), people of other ethnicity had the lowest (5.4%). In contrast, Black Caribbean people had the highest proportion of people with a PCA record for ‘patient unsuitable’ (4.4%) and Chinese people had the lowest (2%) (Table 1).

### ‘Informed Dissent’ PCA findings

In the bivariate analysis (Table 2, Model 1), women were less likely to have a PCA record for ‘informed dissent’ than men [Odds Ratio (OR): 0.81; 95% Confidence Interval (CI): 0.79 to 0.83]. Patients aged 45-59 years and 60 to 74 years were more likely to have a PCA record for ‘informed dissent’ than patients younger than 45 years [OR: 1.34, 95% CI: 1.29 to 1.39 and OR: 1.1, 95% CI: 1.05 to 1.14, respectively]. People with multiple QOF conditions were 2.21 times more likely to have a PCA record for ‘informed dissent’ than people with one QOF condition [95% CI: 2.14 to 2.27]. Patients living in the most deprived quintile were 1.27 times more likely to have a PCA record for ‘informed dissent’ [95% CI: 1.22 to 1.33] than patients living in the least deprived quintile. In multiply adjusted analysis, the association between ‘informed dissent’ and the above covariates followed a similar trend with the exception of age categories where the association was in the opposite direction for people aged 60-74 years and 75 years and above. They were less likely to have a PCA record for informed dissent when compared to those aged 44 years and below [OR: 0.78, 95% CI: 0.75 to 0.81 and OR: 0.63, 95% CI: 0.60 to 0.66] (Table 2, Model 4).

**Table 2.** Logistic regression models showing the association between having a PCA record for ‘informed dissent’ and demographic characteristics.

|  |  | Model 1<br>Covariates assessed separately |  |  | Model 2<br>Adjusted for age and gender |  |  | Model 3<br>Model 2 + multiple QOF conditions |  |  | Model 4<br>Model 3 + area level deprivation |  |  |
| --- | --- | --- | --- | --- | --- | --- | --- | --- | --- | --- | --- | --- | --- |
|  |  | ORs | 95%CI | p | ORs | 95%CI | p | ORs | 95%CI | p | ORs | 95%CI | p |
| **Ethnicity** |  |  |  |  |  |  |  |  |  |  |  |  |  |
|  | White | 1.00 |  |  | 1.00 |  |  | 1.00 |  |  | 1.00 |  |  |
|  | Bangladeshi | 0.8 | 0.63 – 0.99 | **0.05** | 0.79 | 0.62 – 0.98 | **0.04** | 0.74 | 0.58 – 0.92 | **0.01** | 0.69 | 0.55 – 0.87 | **0.002** |
|  | Black African | 0.76 | 0.67 – 0.87 | **<0.001** | 0.73 | 0.63 – 0.83 | **<0.001** | 0.75 | 0.65 – 0.85 | **<0.001** | 0.70 | 0.61 – 0.81 | **<0.001** |
|  | Black Caribbean | 0.75 | 0.66 – 0.86 | **<0.001** | 0.74 | 0.64 – 0.84 | **<0.001** | 0.71 | 0.62 – 0.81 | **<0.001** | 0.67 | 0.58 – 0.76 | **<0.001** |
|  | Black Other | 0.91 | 0.72 – 1.13 | 0.40 | 0.89 | 0.71 – 1.10 | 0.30 | 0.89 | 0.71 – 1.10 | 0.30 | 0.85 | 0.67 – 1.05 | 0.15 |
|  | Chinese | 0.80 | 0.59 – 1.06 | 0.13 | 0.80 | 0.59 – 1.06 | 0.13 | 0.88 | 0.65 – 1.17 | 0.40 | 0.88 | 0.65 – 1.17 | 0.39 |
|  | Indian | 0.80 | 0.71 – 0.89 | **<0.001** | 0.78 | 0.70 – 0.87 | **<0.001** | 0.75 | 0.67 – 0.84 | **<0.001** | 0.74 | 0.66 – 0.83 | **<0.001** |
|  | Mixed | 0.87 | 0.75 – 1.00 | 0.06 | 0.88 | 0.76 – 1.01 | 0.08 | 0.88 | 0.76 – 1.02 | 0.09 | 0.86 | 0.74 – 0.99 | **0.04** |
|  | Other Asian | 0.77 | 0.67 – 0.89 | **0.001** | 0.75 | 0.65 – 0.87 | **<0.001** | 0.75 | 0.65 – 0.87 | **<0.001** | 0.74 | 0.64 – 0.86 | **<0.001** |
|  | Other | 0.66 | 0.55 – 0.79 | **<0.001** | 0.65 | 0.53 – 0.77 | **<0.001** | 0.68 | 0.56 – 0.81 | **<0.001** | 0.66 | 0.55 – 0.80 | **<0.001** |
|  | Pakistani | 1.13 | 1.00 – 1.28 | **0.05** | 1.11 | 0.98 – 1.25 | 0.10 | 1.05 | 0.93 – 1.19 | 0.41 | 1.01 | 0.89 – 1.14 | 0.93 |
| **Gender** |  |  |  |  |  |  |  |  |  |  |  |  |  |
|  | Men | 1.00 |  |  | 1.00 |  |  | 1.00 |  |  | 1.00 |  |  |
|  | Women | 0.81 | 0.79 – 0.83 | **<0.001** | 0.81 | 0.79 – 0.84 | **<0.001** | 0.81 | 0.79 – 0.83 | **<0.001** | 0.81 | 0.79 – 0.83 | **<0.001** |
| **Age** |  |  |  |  |  |  |  |  |  |  |  |  |  |
|  | *<45 | 1.00 |  |  | 1.00 |  |  | 1.00 |  |  | 1.00 |  |  |
|  | 45-59 | 1.34 | 1.29 – 1.39 | **<0.001** | 1.34 | 1.286 – 1.39 | **<0.001** | 1.13 | 1.08 – 1.17 | **<0.001** | 1.13 | 1.09 – 1.18 | **<0.001** |
|  | 60-74 | 1.10 | 1.05 – 1.14 | **<0.001** | 1.08 | 1.038 – 1.13 | **<0.001** | 0.77 | 0.74 – 0.80 | **<0.001** | 0.78 | 0.75 – 0.81 | **<0.001** |
|  | 75+ | 1.00 | 0.95 – 1.04 | 0.85 | 0.99 | 0.948 – 1.04 | 0.68 | 0.62 | 0.59 – 0.64 | **<0.001** | 0.63 | 0.60 – 0.66 | **<0.001** |
| **Multiple QOF conditions** |  |  |  |  |  |  |  |  |  |  |  |  |  |
|  | 1 condition | 1.00 |  |  |  |  |  | 1.00 |  |  | 1.00 |  |  |
|  | 2+ conditions | 2.21 | 2.14 – 2.27 | **<0.001** |  |  |  | 2.55 | 2.47 – 2.63 | **<0.001** | 2.52 | 2.44 – 2.60 | **<0.001** |
| **Area-level Deprivation** |  |  |  |  |  |  |  |  |  |  |  |  |  |
|  | Least deprived Quintile 1 | 1.00 |  |  |  |  |  |  |  |  | 1.00 |  |  |
|  | Quintile 2 | 1.01 | 0.97 – 1.06 | 0.59 |  |  |  |  |  |  | 1.00 | 0.96 – 1.05 | 0.86 |
|  | Quintile 3 | 1.07 | 1.02 – 1.12 | **0.01** |  |  |  |  |  |  | 1.05 | 1.00 – 1.10 | 0.05 |
|  | Quintile 4 | 1.13 | 1.08 – 1.18 | **<0.001** |  |  |  |  |  |  | 1.10 | 1.05 – 1.15 | **<0.001** |
|  | Quintile 5 | 1.27 | 1.22 – 1.33 | **<0.001** |  |  |  |  |  |  | 1.21 | 1.15 – 1.26 | **<0.001** |

Observations = 25046
R<sup>2</sup> Tjur - Model 1 when covariates assessed separately: Ethnicity=0; Gender=0.001; Age categories=0.001, Area-level deprivation=0.001; Multiple QOF conditions=0.012.
R<sup>2</sup> Tjur - Model 2 = 0.002; Model 3&4=0.017

In the unadjusted and fully adjusted models, there were lower odds of having a PCA record for ‘informed dissent’ for Bangladeshi, Black African, Black Caribbean, Indian, other Asian and other ethnic group people (Table 2). People of Pakistani ethnicity were more likely to have a PCA record for ‘informed dissent than people of white ethnicity [OR: 1.13, 95% CI: 1.00 to 1.28] (Table 2, Model 1). This association was no longer evident after adjusting for the sociodemographic characteristics in the other models. Ethnic differences for ‘informed dissent’ were similar for men and women (Supplementary Table 4).

### ‘Patient Unsuitable’ PCA findings

In bivariate analysis, older people and women were more likely to have a PCA record for ‘patient unsuitable’ (Table 3). Those with multiple QOF conditions were 4.69 times more likely to have a PCA record for ‘patient unsuitable’ than those with one condition [95% CI: 4.48 to 4.91]. Also, people living in the fifth most deprived quintile were 1.25 times more likely to have a PCA record for ‘patient unsuitable’ than those living in the least deprived quintile [95% CI: 1.04 to 1.19]. A similar pattern was observed in the multiply adjusted models for these covariates, albeit with some attenuation to the effect sizes (Model 2, 3, 4).

**Table 3.**
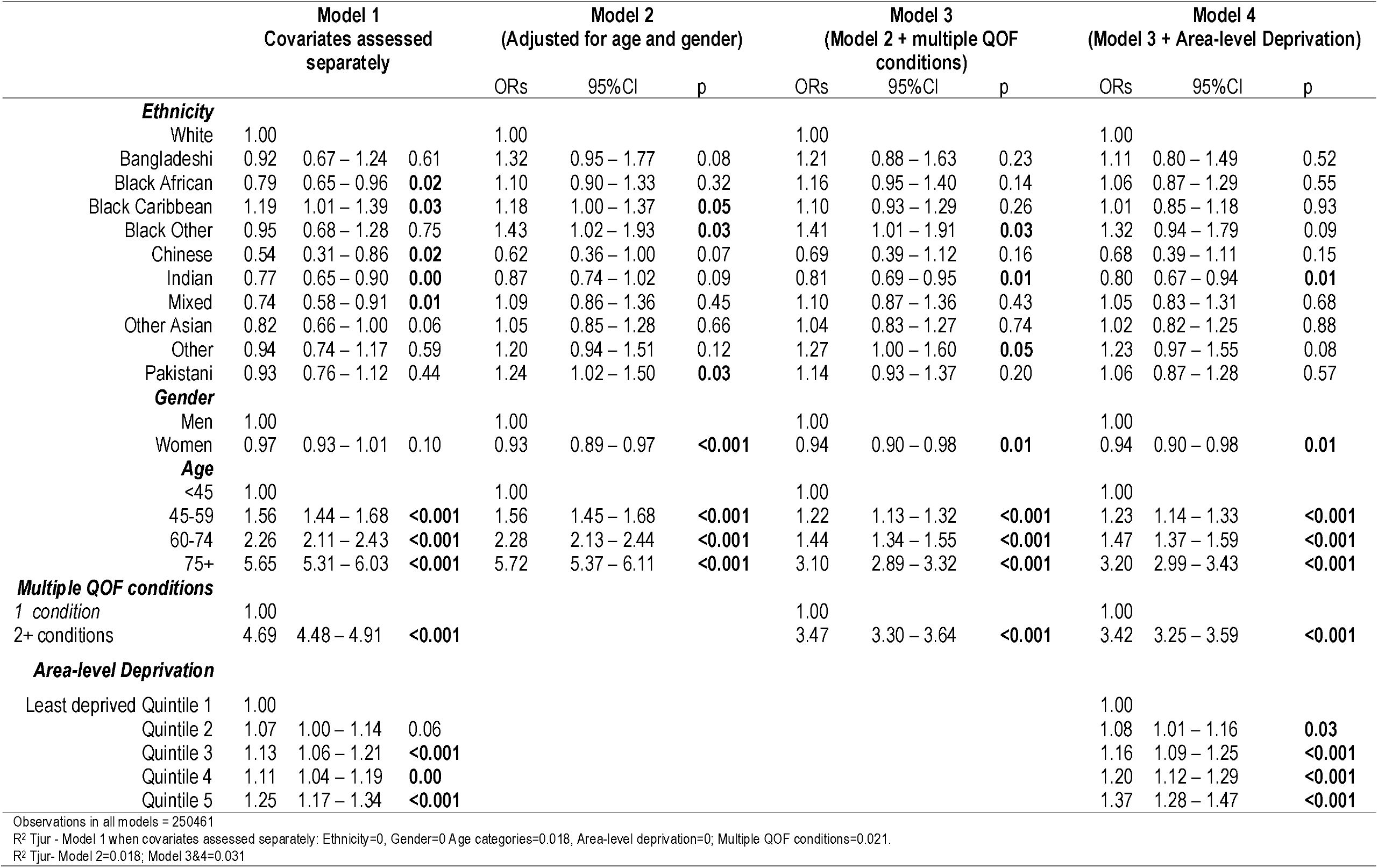
Logistic regression models showing the association between having a PCA record for ‘patient unsuitable’ and demographic characteristics.

Black Caribbean, Black other, Pakistani and other ethnic group patients had higher odds than white patients in the partially adjusted models (Table 3). These inequalities were no longer evident in the final model where we adjusted for age, gender, multiple QOF conditions and area-level deprivation (Table 3, Model 4). Relative to white people, Indian people had lower odds of receiving a PCA record for ‘patient unsuitable’ in the unadjusted model [OR:0.77, 95% CI 0.65 to 0.90] and in the fully adjusted model [OR: 0.81, 95%CI: 0.69 to 0.95] (Table 3, Model 1, 3). Ethnic differences in having a PCA record for ‘patient unsuitable’ were similar for men and women (Supplementary Table 5).

## DISCUSSION

In this study we assessed the patterns of PCA reporting for ‘informed dissent’ and ‘patient unsuitable’, how they varied by ethnic group and whether these patterns could be explained by age, gender, multiple QOF conditions and area-level deprivation. The associations between ethnicity and the two PCA reasons were in opposite directions. Most minoritised ethnic group people were less likely to have a PCA record for ‘informed dissent’. This association was significant for people of Bangladeshi, Black African, Black Caribbean, Indian, mixed, other Asian and other ethnicity after accounting for age, gender, multiple QOF conditions and area-level deprivation. The observed ethnic inequities in PCA reporting for ‘patient unsuitable’ among people of Black Caribbean, Black other, Pakistani, and other ethnicity could be explained by multiple QOF conditions and/or area level deprivation. Only people of Indian ethnicity were significantly less likely than white people to have a PCA record for ‘patient unsuitable’.

### Findings in context

The few studies that explored ethnicity and PCA record focused on exclusion from quality indicators for a particular condition or aggregated minoritised ethnic groups. ^1, 4, 7, 14^ Our study aligns with a previous ecological analysis showing lower levels of ‘informed dissent’ exceptions among people from minoritised ethnic groups ^7^ and builds on their evidence by disaggregating ethnicity looking at PCA reasons separately. Our study also aligns with previous work showing higher rates of PCA for ‘informed dissent’ than for ‘patient unsuitable’ and higher likelihood of having a PCA record for these two reasons with increasing age, area-level deprivation and multimorbidity. ^6^ In their study, women had higher odds of having a PCA record for being unsuitable, a finding which was contrary to our observations. Their conditions of interest included epilepsy, stroke, learning disability and hypothyroidism, which we did not include because these conditions did not have PCA codes for both ‘informed dissent’ and ‘patient unsuitable’.

### Possible mechanisms

Our finding that some minoritised ethnic groups are less likely to have a PCA record for ‘informed dissent’ when compared to the white majority ethnic group raises a number of questions regarding the possible underlying mechanisms. Patients who do not agree to treatment or medical investigation can be considered to be ‘non-compliant’ or ‘non-adherent’.^15^ Racialised explanations such as cultural and attitudinal differences have been provided to describe why some minoritised ethnic group people refuse health services (e.g. clinical examinations, organ transplantation, blood transfusion, antenatal screening and immunisations) resulting in differential health outcomes and healthcare delivery. ^16^ Notwithstanding that such explanations ignore upstream processes which can influence how patients from minoritised ethnic groups make decisions regarding their health, including institutional racism which has fostered mistrust of institutions, ^17^ our findings suggest that they broadly accept and are compliant with the incentivised treatment.^7^

Viewed from this perspective, this finding counters dominant narratives that minoritised ethnic group people often refuse treatment or do not follow health recommendations due to attitudinal differences, cultural and religious beliefs. ^16^

However, provision of ‘informed dissent’ allows GPs to respect patient choice, ^18^ which is seen as part of a general shift towards empowerment ^19^ and increasingly recognised as crucial for preventing illness, maintaining health, and improving healthcare provision and patient experience. ^20^ This perspective represents a shift from a top-down approach to healthcare provision towards an approach that is patient-centred where healthcare providers and patients build a sustainable partnership that can lead to mutual agreement about treatment. ^15, 21^ The lower likelihood of having a PCA record for ‘informed dissent’ for some minoritised ethnic group people could indicate higher levels of disempowerment.

The extant literature suggests that minoritised ethnic group people are more likely to experience a subjective sense of disempowerment due to, for example, cultural insensitivity and discriminatory practices within and beyond the healthcare setting.^22^ Lawrence and colleagues explored ethnic differences in the long-term experiences of living with psychosis and navigating mental health services. ^23^ They highlight how negative expectations and experiences of these services are compounded over time, *creating a vicious cycle of disempowerment and mistrust that manifests for many in resistance to – or at the best passive acceptance of – intervention by mental health services*. ^23^ These findings illuminate the complex relationship between (dis)empowerment and patient dissent and/or assent for minoritised ethnic group people. Future studies that consider the doctor-patient relationship are required to unpack this finding further.

We found that people of Black African, Black other, Pakistani, and other ethnicity were more likely to have a PCA record for ‘patient unsuitable’ compared to white people. These inequities were explained by multiple QOF conditions and area-level deprivation; minoritised ethnic group people are more likely than their white counterparts to be unsuitable for treatment by virtue of the complex intersection of MLTCs with deprivation. Area-level deprivation and multimorbidity are inextricably linked and many, but not all, minoritised ethnic group people have poorer health outcomes that stem from socioeconomic inequities driven by structural, institutional and interpersonal racism and discrimination. ^24-26^ Recent studies also suggest that some minoritised ethnic groups have a higher prevalence of complex multimorbidity. ^27^ which increases the likelihood of polypharmacy. This, in turn, increases susceptibility to inappropriate use of medications and adverse drug reactions. ^28^ The link between multiple conditions and patient unsuitability has also been articulated by Simpson and colleagues who suggest that patients with comorbidities are at an increased risk of being excluded from achievement of clinical targets because they are more likely to be intolerant to certain therapies or multiple treatments which can result in adverse events. ^29^

### Strengths and limitations

The large sample size made available via CPRD Aurum allowed for the disaggregation of the minoritised ethnic group population. However, we were unable to disaggregate the white ethnic group and acknowledge that this is also a diverse population with groups such as the Gypsy, Roma, and Irish Travellers who have poor health outcomes resulting from discrimination and multiple disadvantage. ^24, 30^

In this study we focused on patient-level factors and their impact on PCA patterns. However, practice-level factors such as the number of registered patients, number of GPs, practice deprivation, previous QOF performance, or Personal Medical Services Contract also impact on the rates of PCA recorded. ^6, 7^ Practices located in more deprived areas have a higher tendency to exclude patients for all reasons and for informed dissent. ^7^ Follow up studies are required to assess the association between ethnicity and PCAs and the extent to which key practice-level factors impact on the magnitude and direction of associations observed in this study.

We counted QOF conditions at baseline and did not consider onset or remission of conditions or changes to QOF rules during follow-up. The conditions we included are chronic and changes would have applied across all ethnic groups so it is not clear that this would have introduced bias but we did not test this directly.

## CONCLUSIONS

Some view PCAs as a marker of quality because GPs who practice patient-centred, evidence-based care will inevitably have higher rates of PCAs. ^31^ However, others are concerned from a public health perspective that applying PCAs can lead to a focus on patients more likely to meet targets and a corresponding reduction in the care quality given to exempted patients, thereby, leading to an increase in health inequities ^32-34^ Further, exclusions from pay-for-performance schemes means that we are less likely to have high quality intelligence to guide improved healthcare. Our finding concerning the lower levels of PCA reporting for ‘informed dissent’ among seven of the ten minoritised ethnic groups have not only countered the prevailing narratives that suggest that minoritised ethnic group people refuse medical treatment, but it also illuminates the complex relationship between (dis)empowerment and ‘informed dissent’ which requires further exploration. Additionally, given that PCA recording is closely monitored to reduce misuse and ensure equitable care, it is important to consider that reducing rates of PCA recording for ‘informed dissent’ might come at a cost of disempowerment.

We observed inequities in PCA reporting for ‘patient unsuitability’ among Black African, Black Other, Pakistani, and Other ethnic group people which were attenuated when we accounted for multiple QOF conditions and area level deprivation. These groups may have unmet need and our analysis can inform strategies to ensure all who are eligible receive recommended standards of care. Given that exempting patients from performance targets is associated with poor disease management, ^5^ as well as poor survival, ^6^ this finding provides insight into the mechanisms driving ethnic inequities in care that should be addressed in the interest of preventing poor health outcomes.

## Data Availability

This study uses routinely collected individual patient data which can be obtained from Clinical Research Practice Datalink subject to protocol approval via the CPRD Research Data Governance Process. Although these data are anonymised, they are considered sensitive data in the UK by the Data Protection Act and, therefore, cannot be shared publicly. Information about applying to use CPRD data can be found at https://www.cprd.com/data-access.

---

**Supplementary Table 1.**
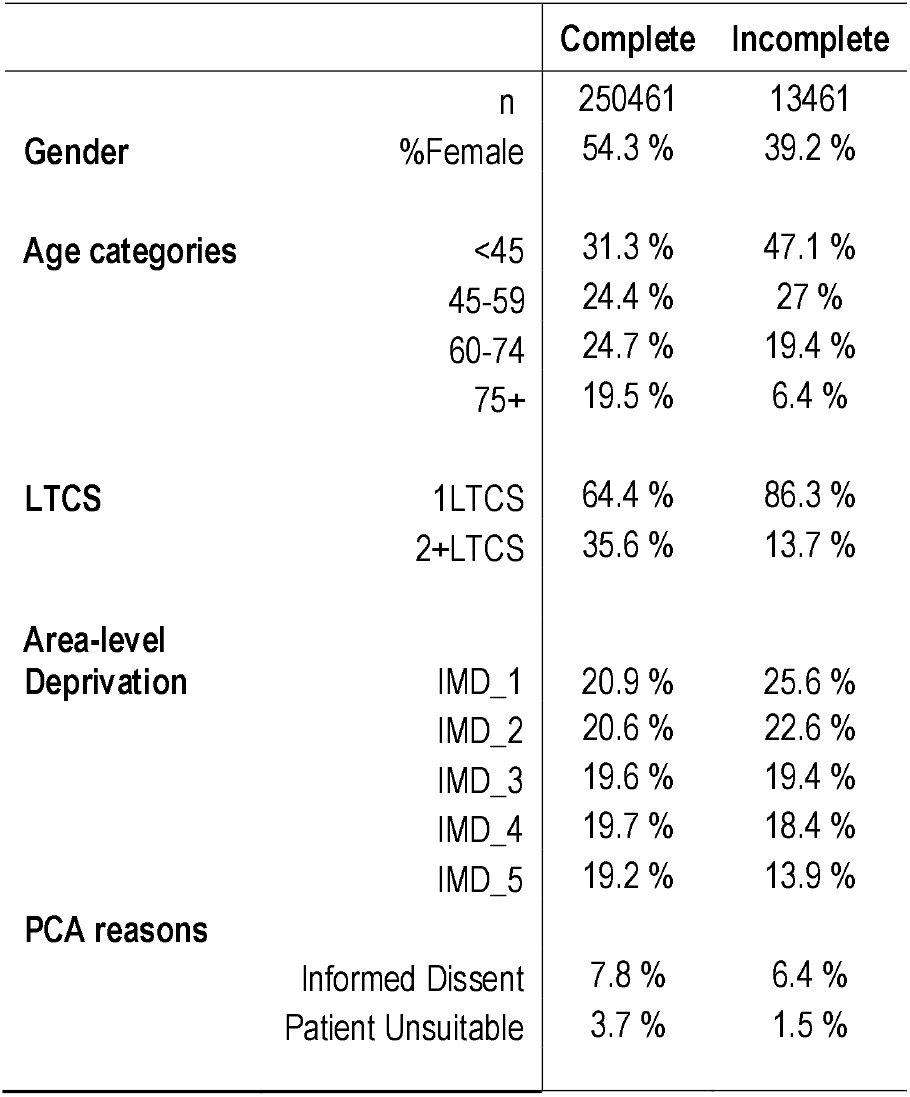
Missing ethnicity data.

**Supplementary Table 2.** Number of patients with each QOF condition.

| QOF condition | n |
| --- | --- |
| Asthma | 81028 |
| Atrial Fibrillation | 11859 |
| Cancer | 21719 |
| Coronary Heart Disease | 21075 |
| Chronic Obstructive Pulmonary Disease | 11425 |
| Dementia | 4887 |
| Depression | 90618 |
| Diabetes | 30603 |
| Heart Failure | 4530 |
| Hypertension | 91403 |
| Rheumatoid Arthritis | 3870 |
| Severe Mental Illness | 6409 |
| At least one QOF condition | 250461 |

**Supplementary Table 3.** Demographic characteristics of sample.

|  |  | Ethnicity |  |  |  |  |  |  |  |  |  |  |  |
| --- | --- | --- | --- | --- | --- | --- | --- | --- | --- | --- | --- | --- | --- |
|  |  | Bangladeshi | Black African | Black Caribbean | Black Other | Chinese | Indian | Mixed | Other Asian | Other | Pakistani | White | Total |
| Gender | Men | 629<br>(50.2 %) | 1705<br>(46.7 %) | 1626<br>(43.5 %) | 534<br>(46 %) | 323<br>(43.6 %) | 2608<br>(49.1 %) | 1266<br>(43.7 %) | 1487<br>(49 %) | 1082<br>(50.2 %) | 1615<br>(50.1 %) | 101482<br>(45.4 %) | **114357**<br>**(45.7 %)** |
|  | Women | 624<br>(49.8 %) | 1947<br>(53.3 %) | 2111<br>(56.5 %) | 628<br>(54 %) | 418<br>(56.4 %) | 2701<br>(50.9 %) | 1634<br>(56.3 %) | 1548<br>(51 %) | 1072<br>(49.8 %) | 1606<br>(49.9 %) | 121815<br>(54.6 %) | **136104**<br>**(54.3 %)** |
| Age categories | <45 | 638<br>(50.9 %) | 1360<br>(37.2 %) | 969<br>(25.9 %) | 576<br>(49.6 %) | 265<br>(35.8 %) | 1673<br>(31.5 %) | 1594<br>(55 %) | 1129<br>(37.2 %) | 884<br>(41 %) | 1405<br>(43.6 %) | 67974<br>(30.4 %) | **78467**<br>**(31.3 %)** |
|  | 45-59 | 338<br>(27 %) | 1457<br>(39.9 %) | 1196<br>(32 %) | 392<br>(33.7 %) | 179<br>(24.2 %) | 1452<br>(27.3 %) | 732<br>(25.2 %) | 911<br>(30 %) | 642<br>(29.8 %) | 907<br>(28.2 %) | 52915<br>(23.7 %) | **61121**<br>**(24.4 %)** |
|  | 60-74 | 182<br>(14.5 %) | 610<br>(16.7 %) | 749<br>(20 %) | 123<br>(10.6 %) | 204<br>(27.5 %) | 1478<br>(27.8 %) | 372<br>(12.8 %) | 730<br>(24.1 %) | 412<br>(19.1 %) | 644<br>(20 %) | 56446<br>(25.3 %) | **61950**<br>**(24.7 %)** |
|  | 75+ | 95<br>(7.6 %) | 225<br>(6.2 %) | 823<br>(22 %) | 71<br>(6.1 %) | 93<br>(12.6 %) | 706<br>(13.3 %) | 202<br>(7 %) | 265<br>(8.7 %) | 216<br>(10 %) | 265<br>(8.2 %) | 45962<br>(20.6 %) | **48923**<br>**(19.5 %)** |
| Area-level Deprivation | Quintile 1 | 65<br>(5.2 %) | 124<br>(3.4 %) | 142<br>(3.8 %) | 63<br>(5.4 %) | 156<br>(21.1 %) | 863<br>(16.3 %) | 336<br>(11.6 %) | 420<br>(13.8 %) | 287<br>(13.3 %) | 227<br>(7 %) | 49639<br>(22.2 %) | **52322**<br>**(20.9 %)** |
|  | Quintile 2 | 81<br>(6.5 %) | 239<br>(6.5 %) | 244<br>(6.5 %) | 125<br>(10.8 %) | 143<br>(19.3 %) | 871<br>(16.4 %) | 378<br>(13 %) | 526<br>(17.3 %) | 315<br>(14.6 %) | 301<br>(9.3 %) | 48366<br>(21.7 %) | **51589**<br>**(20.6 %)** |
|  | Quintile 3 | 127<br>(10.1 %) | 484<br>(13.3 %) | 587<br>(15.7 %) | 156<br>(13.4 %) | 147<br>(19.8 %) | 1198<br>(22.6 %) | 526<br>(18.1 %) | 663<br>(21.8 %) | 403<br>(18.7 %) | 500<br>(15.5 %) | 44359<br>(19.9 %) | **49150**<br>**(19.6 %)** |
|  | Quintile 4 | 349 | 1131 | 1168 | 336 | 173 | 1356 | 769 | 856 | 578 | 877 | 41781 | **49374** |
|  |  | (27.9 %) | (31 %) | (31.3 %) | (28.9 %) | (23.3 %) | (25.5 %) | (26.5 %) | (28.2 %) | (26.8 %) | (27.2 %) | (18.7 %) | **(19.7 %)** |
|  | Quintile 5 | 631<br>(50.4 %) | 1674<br>(45.8 %) | 1596<br>(42.7 %) | 482<br>(41.5 %) | 122<br>(16.5 %) | 1021<br>(19.2 %) | 891<br>(30.7 %) | 570<br>(18.8 %) | 571<br>(26.5 %) | 1316<br>(40.9 %) | 39152<br>(17.5 %) | **48026**<br>**(19.2 %)** |
| **Number of QOF conditions** | 1 QOF condition | 846<br>(67.5 %) | 2702<br>(74 %) | 2207<br>(59.1 %) | 859<br>(73.9 %) | 562<br>(75.8 %) | 3283<br>(61.8 %) | 2175<br>(75 %) | 2088<br>(68.8 %) | 1612<br>(74.8 %) | 2102<br>(65.3 %) | 142985<br>(64 %) | **161421**<br>**(64.4 %)** |
|  | 2+ QOF conditions | 407<br>(32.5 %) | 950<br>(26 %) | 1530<br>(40.9 %) | 303<br>(26.1 %) | 179<br>(24.2 %) | 2026<br>(38.2 %) | 725<br>(25 %) | 947<br>(31.2 %) | 542<br>(25.2 %) | 1119<br>(34.7 %) | 80312<br>(36 %) | **89040**<br>**(35.6 %)** |
| **PCA record for 'Informed dissent'** | Present | 1172<br>(93.5 %) | 3425<br>(93.8 %) | 3508<br>(93.9 %) | 1077<br>(92.7 %) | 693<br>(93.5 %) | 4965<br>(93.5 %) | 2696<br>(93 %) | 2844<br>(93.7 %) | 2037<br>(94.6 %) | 2933<br>(91.1 %) | 205458<br>(92 %) | **230808**<br>**(92.2 %)** |
|  | Absent | 81<br>(6.5 %) | 227<br>(6.2 %) | 229<br>(6.1 %) | 85<br>(7.3 %) | 48<br>(6.5 %) | 344<br>(6.5 %) | 204<br>(7 %) | 191<br>(6.3 %) | 117<br>(5.4 %) | 288<br>(8.9 %) | 17839<br>(8 %) | **19653**<br>**(7.8 %)** |
| **PCA record for 'Patient Unsuitable'** | Present | 1210<br>(96.6 %) | 3544<br>(97 %) | 3573<br>(95.6 %) | 1121<br>(96.5 %) | 726<br>(98 %) | 5156<br>(97.1 %) | 2820<br>(97.2 %) | 2942<br>(96.9 %) | 2079<br>(96.5 %) | 3110<br>(96.6 %) | 215023<br>(96.3 %) | **241304**<br>**(96.3 %)** |
|  | Absent | 43<br>(3.4 %) | 108<br>(3 %) | 164<br>(4.4 %) | 41<br>(3.5 %) | 15<br>(2 %) | 153<br>(2.9 %) | 80<br>(2.8 %) | 93<br>(3.1 %) | 75<br>(3.5 %) | 111<br>(3.4 %) | 8274<br>(3.7 %) | **9157**<br>**(3.7 %)** |
|  | **Total** | **1253**<br>**(100 %)** | **3652**<br>**(100 %)** | **3737**<br>**(100 %)** | **1162**<br>**(100 %)** | **741**<br>**(100 %)** | **5309**<br>**(100 %)** | **2900**<br>**(100 %)** | **3035**<br>**(100 %)** | **2154**<br>**(100 %)** | **3221**<br>**(100 %)** | **223297**<br>**(100 %)** | **250461**<br>**(100 %)** |

**Supplementary Table 4.** Logistic regression models showing the association between having a PCA record for ‘informed dissent’ and demographic characteristics.

|  | **Men** |  |  | **Women** |  |  |
| --- | --- | --- | --- | --- | --- | --- |
|  | ORs | 95%CI | p | ORs | 95%CI | p |
| ***Ethnicity*** |  |  |  |  |  |  |
| White | 1.00 |  |  | 1.00 |  |  |
| Bangladeshi | 0.62 | 0.444 – 0.844 | **0.004** | 0.78 | 0.557 – 1.067 | 0.14 |
| Black African | 0.72 | 0.591 – 0.860 | **<0.001** | 0.69 | 0.562 – 0.841 | **<0.001** |
| Black Caribbean | 0.72 | 0.595 – 0.873 | **0.001** | 0.62 | 0.509 – 0.748 | **<0.001** |
| Black Other | 0.91 | 0.665 – 1.227 | 0.566 | 0.78 | 0.555 – 1.071 | 0.14 |
| Chinese | 1.08 | 0.717 – 1.570 | 0.689 | 0.70 | 0.430 – 1.068 | 0.12 |
| Indian | 0.81 | 0.695 – 0.931 | **0.004** | 0.67 | 0.560 – 0.790 | **<0.001** |
| Mixed | 0.99 | 0.810 – 1.198 | 0.915 | 0.74 | 0.591 – 0.911 | **0.01** |
| Other Asian | 0.83 | 0.677 – 0.996 | 0.052 | 0.64 | 0.506 – 0.808 | **<0.001** |
| Other | 0.61 | 0.463 – 0.781 | **<0.001** | 0.73 | 0.550 – 0.949 | **0.02** |
| Pakistani | 0.95 | 0.801 – 1.129 | 0.594 | 1.07 | 0.892 – 1.275 | 0.46 |
| ***Age*** |  |  |  |  |  |  |
| *<45 | 1.00 |  |  | 1.00 |  |  |
| 45-59 | 1.19 | 1.126 – 1.257 | **<0.001** | 1.08 | 1.021 – 1.144 | **0.01** |
| 60-74 | 0.76 | 0.715 – 0.806 | **<0.001** | 0.81 | 0.762 – 0.860 | **<0.001** |
| 75+ | 0.56 | 0.517 – 0.597 | **<0.001** | 0.69 | 0.651 – 0.740 | **<0.001** |
| ***Multiple QOF conditions*** |  |  |  |  |  |  |
| *1 LTC | 1.00 |  |  | 1.00 |  |  |
| 2+ LTCS | 2.28 | 2.181 – 2.390 | **<0.001** | 2.79 | 2.670 – 2.921 | **<0.001** |
| ***Area-level Deprivation (IMD)*** |  |  |  |  |  |  |
| *Quintile 1 | 1.00 |  |  | 1.00 |  |  |
| Quintile 2 | 1.01 | 0.945 – 1.079 | 0.78 | 1.00 | 0.932 – 1.066 | 0.93 |
| Quintile 3 | 1.04 | 0.974 – 1.113 | 0.241 | 1.05 | 0.984 – 1.125 | 0.14 |
| Quintile 4 | 1.09 | 1.021 – 1.166 | **0.01** | 1.11 | 1.034 – 1.182 | **0.00** |
| Quintile 5 | 1.18 | 1.100 – 1.255 | **<0.001** | 1.23 | 1.151 – 1.314 | **<0.001** |
| Observations |  |  | 114357 |  |  | 136104 |
| R<sup>2</sup> Tjur |  |  | 0.016 |  |  | 0.018 |

**Supplementary Table 5.** Logistic regression models showing the association between having a PCA record for ‘Patient Unsuitable’ and demographic characteristics.

|  | **Men** |  |  | **Women** |  |  |
| --- | --- | --- | --- | --- | --- | --- |
|  | ORs | 95%CI | p | ORs | 95%CI | p |
| ***Ethnicity*** |  |  |  |  |  |  |
| White | 1.00 |  |  | 1.00 |  |  |
| Bangladeshi | 1.28 | 0.842 – 1.857 | 0.22 | 0.89 | 0.515 – 1.424 | 0.65 |
| Black African | 0.96 | 0.718 – 1.264 | 0.80 | 1.16 | 0.876 – 1.514 | 0.28 |
| Black Caribbean | 1.16 | 0.912 – 1.442 | 0.22 | 0.90 | 0.709 – 1.124 | 0.36 |
| Black Other | 1.43 | 0.896 – 2.150 | 0.11 | 1.21 | 0.735 – 1.879 | 0.42 |
| Chinese | 0.60 | 0.234 – 1.228 | 0.21 | 0.78 | 0.368 – 1.428 | 0.46 |
| Indian | 0.87 | 0.691 – 1.077 | 0.21 | 0.72 | 0.559 – 0.916 | **0.01** |
| Mixed | 1.02 | 0.715 – 1.395 | 0.93 | 1.09 | 0.787 – 1.467 | 0.59 |
| Other Asian | 0.82 | 0.580 – 1.109 | 0.21 | 1.23 | 0.921 – 1.610 | 0.14 |
| Other | 1.29 | 0.921 – 1.745 | 0.12 | 1.15 | 0.793 – 1.601 | 0.44 |
| Pakistani | 1.12 | 0.856 – 1.432 | 0.40 | 0.99 | 0.729 – 1.322 | 0.97 |
| ***Age*** |  |  |  |  |  |  |
| <45 | 1.00 |  |  | 1.00 |  |  |
| 45-59 | 1.25 | 1.126 – 1.392 | **<0.001** | 1.19 | 1.066 – 1.331 | **0.00** |
| 60-74 | 1.32 | 1.186 – 1.460 | **<0.001** | 1.63 | 1.474 – 1.813 | **<0.001** |
| 75+ | 2.52 | 2.279 – 2.795 | **<0.001** | 3.87 | 3.525 – 4.264 | **<0.001** |
| ***Multiple QOF conditions*** |  |  |  |  |  |  |
| 1 condition | 1.00 |  |  | 1.00 |  |  |
| 2+ conditions | 3.32 | 3.093 – 3.574 | **<0.001** | 3.54 | 3.311 – 3.793 | **<0.001** |
| ***Area-level deprivation (IMD)*** |  |  |  |  |  |  |
| Quintile 1 | 1.00 |  |  | 1.00 |  |  |
| Quintile 2 | 1.12 | 1.012 – 1.237 | **0.03** | 1.05 | 0.954 – 1.148 | 0.34 |
| Quintile 3 | 1.20 | 1.083 – 1.324 | **<0.001** | 1.13 | 1.032 – 1.242 | **0.01** |
| Quintile 4 | 1.20 | 1.084 – 1.329 | **<0.001** | 1.19 | 1.082 – 1.304 | **<0.001** |
| Quintile 5 | 1.48 | 1.339 – 1.633 | **<0.001** | 1.27 | 1.157 – 1.395 | **<0.001** |
| Observations |  |  | 114357 |  |  | 136104 |
| R<sup>2</sup> Tjur |  |  | 0.025 |  |  | 0.038 |

## Notes

### Competing Interest Statement

MS and JH are employed by The Health Foundation. The authors have no competing interest to declare.

### Funding Statement

This work is funded by The Health Foundation (AIMS 1874695).

### Author Declarations

This study is part of a wider project investigating ethnic inequities in health care use and care quality among people with multiple conditions. Ethics committee/IRB of University of Sussex gave ethical approval for this project. The study was reviewed for ethical and methods content and approved by the CPRD team (eRAP protocol number 21_000333).

